# Fifteen Days in December: Capture and Analysis of Omicron-Related Travel Restrictions

**DOI:** 10.1101/2022.01.26.22269910

**Authors:** Jordan Schermerhorn, Alaina Case, Ellie Graeden, Justin Kerr, Mackenzie Moore, Siobhan Robinson-Marshall, Trae Wallace, Emily Woodrow, Rebecca Katz

**Affiliations:** Center for Global Health Science and Security, Georgetown University, Washington, DC, United States; Talus Analytics, 1855 57^th^ Court, Suite 200, Boulder, CO 80540

## Abstract

Following the identification of the Omicron variant of the SARS-CoV-2 virus in late November 2021, governments worldwide took actions intended to minimize the impact of the new variant within their borders. Despite guidance from the World Health Organization advising a risk-based approach, many rapidly implemented stringent policies focused on travel restrictions. In this paper, we capture 221 national-level travel policies issued during the three weeks following publicization of the Omicron variant. We characterize policies based upon whether they target travelers from specific countries or focus more broadly on enhanced screening, and explore differences in approaches at the regional level. We find that initial reactions almost universally focused on entry bans and flight suspensions from Southern Africa, and that policies continued to target travel from these countries even after community transmission of the Omicron variant was detected elsewhere in the world. While layered testing and quarantine requirements were implemented by some countries later in this three-week period, these enhanced screening policies were rarely the first response. The timing and conditionality of quarantine and testing requirements were not coordinated between countries or regions, creating logistical complications and burdening travelers with costs. Overall, response measures were rarely tied to specific criteria or adapted to match the unique epidemiology of the new variant.

**Summary box:**

- During the initial three-week period following the discovery of the SARS-CoV-2 Omicron variant, nations rushed to implement travel restrictions - often at odds with guidance from the World Health Organization.
- By sourcing and cataloging initial national-level travel restrictions worldwide, we demonstrate how the distribution of entry bans, flight suspensions, quarantine measures, vaccination requirements, and testing protocols evolved in response to emerging information during a period of uncertainty.
- Countries that issued entry bans almost universally targeted the same Southern African countries and continued to do so even after widespread community transmission of the Omicron variant was reported elsewhere in the world.
- Layers of testing and quarantine requirements were added later during the observation period but were rarely the initial response, with the exception of restrictions issued by countries in Africa, where leading with enhanced screening measures was more common.
- Analysis of the disconnect between travel restrictions and transmission patterns that followed emergence of the Omicron variant provides a basis to inform evidence-based control measures for future virus mitigation efforts.

## Introduction

Following the identification of the Omicron variant of the SARS-CoV-2 virus in South Africa on 24 November 2021 and reports of infection in vaccinated individuals, governments worldwide were tasked with the responsibility of navigating political and risk-based decisions to minimize the impact of the new variant within their borders.[1] In spite of World Health Organization (WHO) guidance that advised against travel bans in this circumstance, many countries implemented a rapid series of measures banning entry and suspending flights from Southern African nations.[2] Leaders from Southern Africa issued strong statements criticizing these measures, and framed the targeted restrictions as an inappropriate punishment for strong genomic surveillance and transparent reporting.[3] The COVID-19 Emergency Committee convened under the International Health Regulations ultimately released a report in January 2022 that both praised South Africa for its contributions and reiterated discouragement of blanket travel bans.[4]

Despite some evidence pointing to travel restrictions as useful for reducing exported cases during very early stages of spread, this context quickly was overridden by the near-immediate detection of the new variant far beyond Southern Africa and its high estimated transmissibility.[5] Previous efforts to characterize types of public health travel restrictions have emphasized that the breadth of measures falling under “travel ban” terminology complicates analysis of their effectiveness.[6] However, as nearly all countries had previously implemented cross-border restrictions at some point during the COVID-19 pandemic, they remained a policy tool that could be rapidly implemented with recent precedent.[7]

In this analysis, we capture and explore the short-term global policy response to the discovery of the Omicron variant of concern of COVID-19. By focusing on travel restrictions issued at the national level, we enable future analysis of case trajectories following different policy approaches.

### Capturing travel restriction policy data

We identified travel-related policies issued between 24 November and 15 December 2021 for all WHO member states, territories, and invited observers. Policies issued during this three-week period were categorized based on whether they imposed restrictions on travelers from specific countries, or impacted all inbound passengers; tagged with one or more topical categories based upon what the policy intended to do; and associated with the WHO Regional Office of the issuing jurisdiction. These six Regional Offices include Africa (AFRO), the Eastern Mediterranean (EMRO), Europe (EURO), the Americas (PAHO), South-East Asia (SEARO), and the Western Pacific (WPRO).

Researchers found policies using publicly available sources and manually entered 23 corresponding metadata in an Airtable database.[8] For complex policies that involved multiple categories, coding approaches were aligned through research team meetings. A static copy (PDF or screen capture) of each policy was stored along with links to any associated websites and summary data in the COVID Analysis and Mapping of Policies (COVID-AMP) database, available at covidamp.org.

Sources included websites for government entities, such as Ministries of Health, Transport, and Foreign Affairs; centralized COVID-19 information portals; legislation and presidential decrees; and regulations from bodies such as Civil Aviation Authorities. Informal dissemination mechanisms, such as official social media accounts from government departments and administrators, were also considered valid sources. In these instances, official policy documents were often posted as images. In the absence of any formal policy documents available from official government sources, policy announcements from national airlines and state media sources were included.

Ultimately, this process identified 221 total Omicron response travel policies issued worldwide during the first three weeks after discovery of the variant was announced.

### Restrictions targeted to specific countries

Beginning with a flight suspension issued by the United Kingdom on 25 November 2021, initial quick policy actions by a small group of countries were rapidly emulated around the world.[9] Countries that implemented any flight restrictions or entry bans almost universally targeted the same seven countries in Southern Africa: South Africa, Eswatini, Lesotho, Botswana, Zimbabwe, Mozambique, and Namibia **(Figure 1)**. A second tier of targeted countries included Zambia, Angola, and Malawi, followed by Nigeria and Egypt.

**Figure 1.**
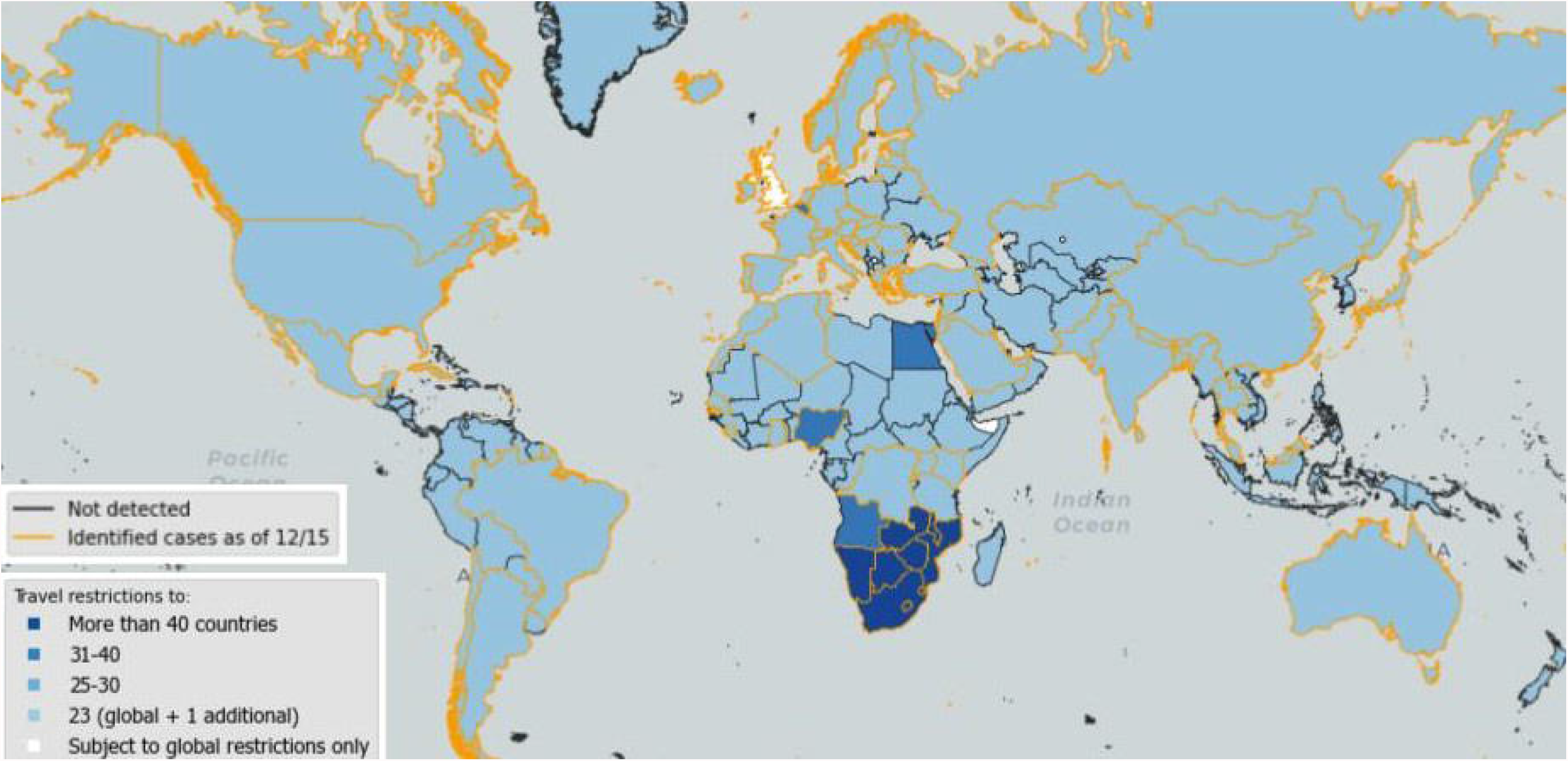
Map illustrating the number of Omicron-related travel restrictions that imposed specific measures against each country, 24 November – 15 December 2021. Countries in which confirmed Omicron cases had been reported as of 15 December are outlined in yellow.

The bulk of these entry bans permitted repatriation of citizens and residents with additional testing and quarantine requirements. However, several specified the prohibition of airport thru- transit for travelers from the same targeted countries, which complicated returns in practice.

In most cases, these restrictions were not tied to specific epidemiological data such as case trajectories, positivity rates, or overall surveillance capabilities of a country. Entry bans targeting Nigeria and Egypt in particular were based on detection of the Omicron variant in people who had recently traveled from those countries, rather than cases reported from within the countries themselves.[10] Several countries later incorporated their Omicron responses into pre-existing travel risk frameworks such as “red lists”, but initial responses often existed outside of these tiered systems.[9]

### Evolution of restrictions over time

Policies enacted immediately after the Omicron announcement tended to focus on entry bans and border closures. Those issued later during the observation period tended to be more nuanced, focusing on testing and other mechanisms to allow for travel but enhance screening at the borders. These were either tailored to the risk levels of incoming passengers – for example, quarantine requirements for travelers arriving from specific countries – or universal, such as adjustments to the time frame in which pre-travel PCR tests must be obtained.[2]

Though many countries that had led with entry bans and flight suspensions later added layers of testing and quarantine for all travelers, their initial targeted restrictions were rarely repealed or rescinded during the first three weeks of response **(Figure 2)**. During this time period, only four countries - Cambodia, Sri Lanka, Israel, and the United Kingdom - lifted their entry bans entirely and replaced them with enhanced screening protocols.

**Figure 2.**
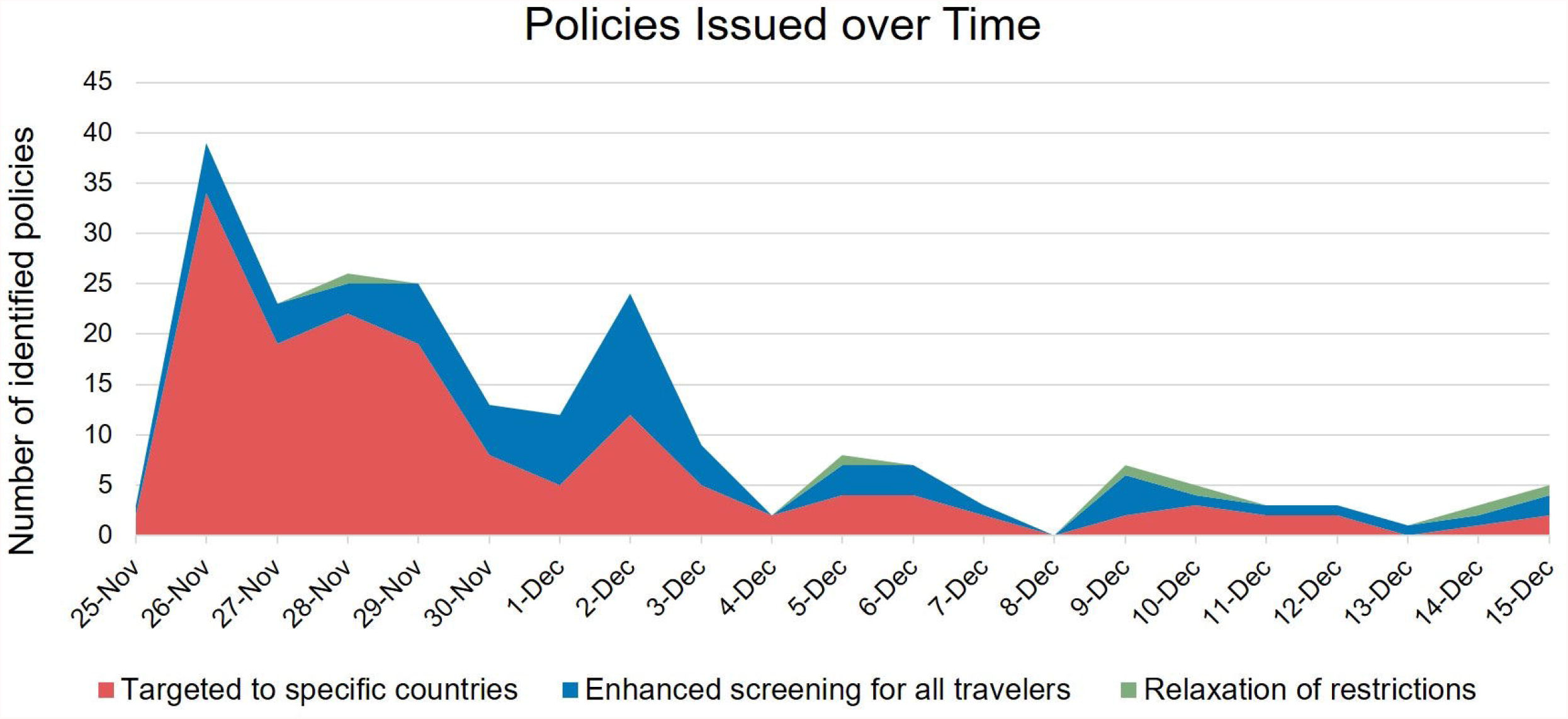
Following an initial burst of policies targeted to specific countries over time, measures became slightly more focused on enhancing screening measures such as testing for all travelers – though initial entry bans were rarely rescinded.

The intended duration of travel restrictions was generally not defined in the policy documentation, and did not include specific criteria for relaxation. Only 15% of restrictions explicitly mentioned an anticipated end date; of these, several were ultimately extended.

### Regional differences in response strategies

Examining policies by the WHO regional office associated with the issuing body, we found substantial differences in the tendency to focus restrictions on specific countries versus updating overall screening protocols.

In press statements, African leaders emphasized the negative impacts of entry bans and the harm of perceived punishment associated with transparent surveillance and reporting.[3] This stance was supported by actions: AFRO was the only region in which more than half of Omicron response policies focused on universal entry requirements, with relatively few specific entry bans **(Figure 3)**.

**Figure 3.**
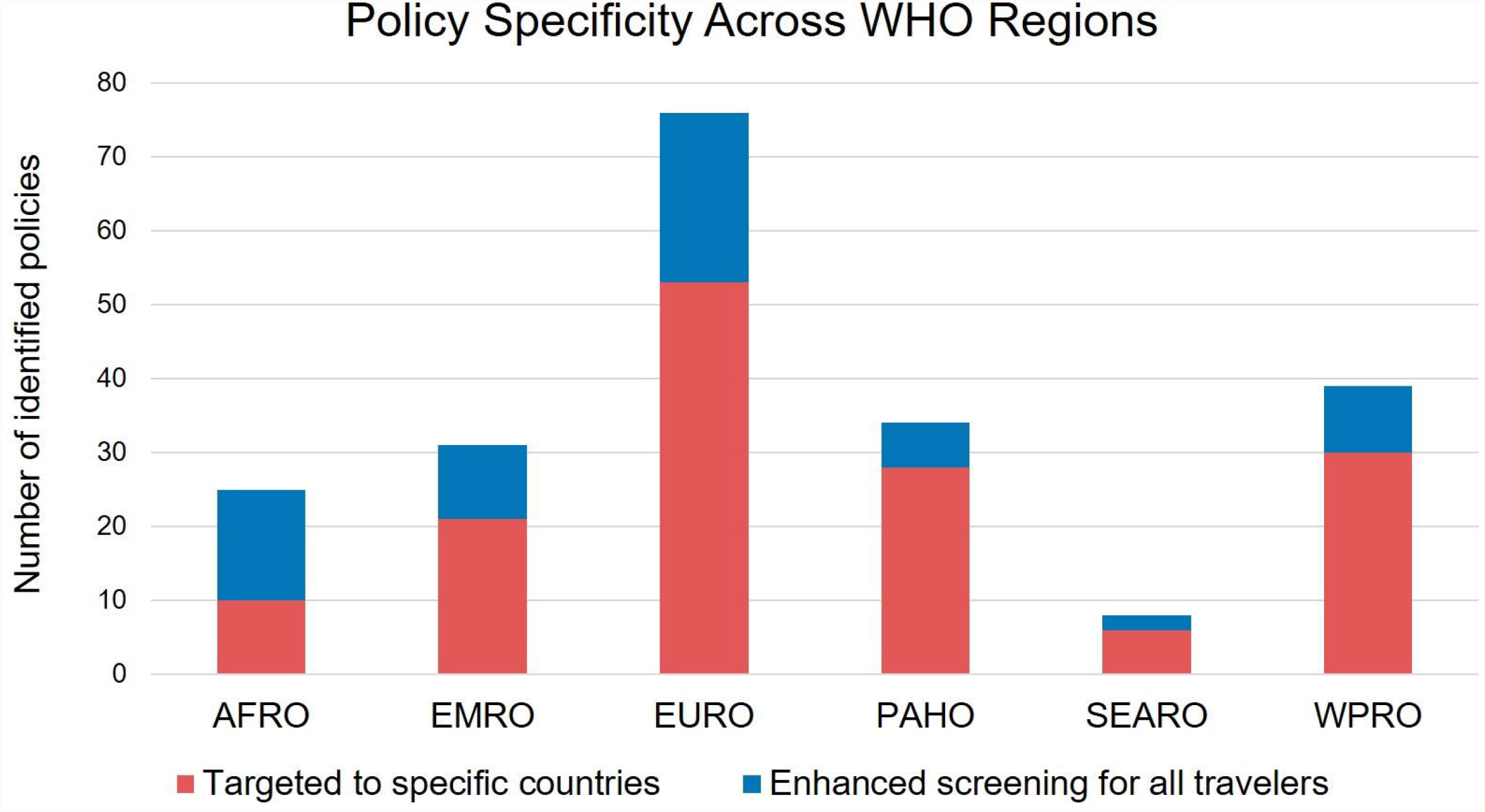
The number of travel restrictions issued by countries in each WHO region between 24 November and 15 December 2021, indicating whether the policies were targeted to specific countries or were relevant to all incoming travelers.

Policies in the EURO region were largely aligned with an “emergency brake” activated by the European Union (EU), which recommended temporary restrictions on all travel into the EU from Botswana, Eswatini, Lesotho, Mozambique, Namibia, South Africa, and Zimbabwe on 26 November 2021. Most countries in the region implemented formal restrictions to enact that recommendation, and several followed the entry ban with additional general precautions.

Beyond the scope of targeted restrictions versus those pertaining to all travelers, we also found qualitative differences in overall approaches to mitigation across WHO regions. Flight suspensions were most commonly issued by EURO and EMRO countries, particularly Gulf nations home to air travel hubs **(Figure 4)**. PAHO and SEARO countries avoided flight suspensions entirely due to the lack of direct travel routes, instead opting to rely on entry bans and screening measures imposed by intermediaries.[11]

**Figure 4.**
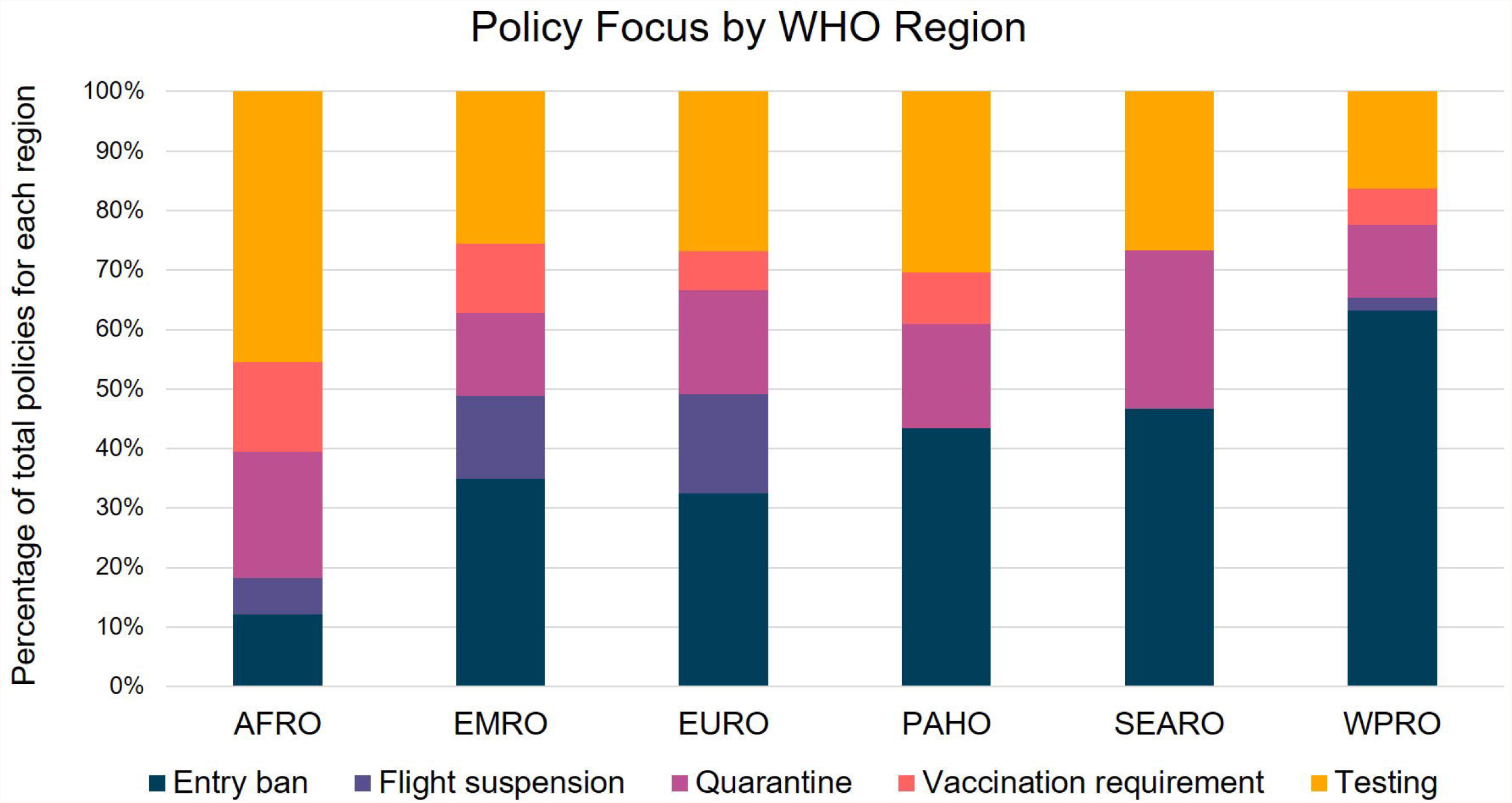
Categorical breakdown of travel restriction policies instituted by countries in each WHO region from 24 November – 15 December 2021.

WPRO countries include many island nations, for whom entry bans may have been easiest to implement and in keeping with more stringent pre-Omicron approaches. Australia, for example, did not permit repatriation of Australian citizens from Southern African countries until 15 December.[12]

AFRO members were the most likely to rely on enhanced testing measures and the least likely to institute entry bans. Notably, many did not hesitate to impose vaccination requirements for entry despite relatively low rates of vaccination within their own populations. Beyond an expression of solidarity with the frequently-targeted Southern African countries, this strategy may have also been influenced in part by suspected already-established community transmission of the Omicron variant throughout much of the continent.

### Variations in enhanced screening measures

Despite adopting the same overall strategies for screening incoming travelers, countries varied in the details of their approaches. Quarantine restrictions ranged from unsupervised quarantine at any place of residence to strict monitoring in designated hotels. Many required travelers to bear quarantine costs, occasionally requiring pre-purchase of quarantine packages including meals.Several quarantine requirements included testing on specified days, though negative tests were not always sufficient to end the quarantine period.[12]

New testing requirements were similarly diverse. Of 81 policies addressing testing, 51 specified that PCR tests were required, while nine explicitly allowed either PCR or rapid antigen tests.

Pre-travel periods for test validity ranged from 24 to 120 hours, with 72 hours being most common. The phrases “before departure”, “before boarding”, and “before arrival” were all used to start the clock. Proactively, some countries in Africa began to require their own citizens to present negative PCR tests before exiting the country, regardless of destination.[12] These requirements may have been intended to preempt or mitigate targeted restrictions from other nations.

Some of these policy updates appear to reflect emerging knowledge about transmission of the Omicron variant, including the shorter average incubation period: Côte d’Ivoire, for instance, shortened its period for pre-departure test validity from 72 to 48 hours on December 9th. 26 policies implemented testing both prior to departure and upon arrival, which may improve identification of passengers who begin testing positive en route. The majority of testing requirements, however, seemed divorced from the epidemiology of transmission and more closely tied to the logistics of receiving results.

### Practical and logistical considerations

Travel restrictions related to the Omicron response posed major logistical challenges worldwide. Flight suspensions delayed the shipment of reagents to researchers and responders on the ground in Southern Africa, counterproductively impacting their ability to learn more about the variant and quickly inform a better global response.[13] While nearly all entry bans permitted repatriation of citizens and residents, the logistics of return were more challenging. Restrictions on intermediate transit severely complicated travel out of Africa, especially from nations reliant on flight paths through Johannesburg. Policies issued by nations home to air travel hubs in Europe and the Middle East had further-reaching consequences, sometimes making repatriation impossible in practice even if it was not directly prohibited. Morocco, for instance, did not ban tourists from leaving the country, but a total suspension of inbound flights meant that planes were not available for later departures.

Testing requirements, even if clearly defined, present travelers with significant challenges, particularly in places with insufficient laboratory capacity or locations with unusually high demand. If countries require a negative test taken within 24 or 48 hours of arrival, obtaining PCR results that quickly can be difficult, and guidance on best practices following a positive result and subsequent rescheduling are often unclear. Moreover, the financial burden of testing and quarantine requirements was nearly always placed on the traveler. That burden can grow significantly in the case of a positive test, leaving travelers to manage housing and access medical care away from home.

Many travel restrictions were also contingent upon traveler vaccination status. Several countries implemented different testing and quarantine requirements for vaccinated and partially unvaccinated travelers – a distinction that might not be best suited to address the immune evasion properties of Omicron. Countries also define “fully vaccinated” differently: Saudi Arabia, for example, began requiring booster doses of mRNA vaccines to achieve full vaccination status.[12] Lack of clarity on what full vaccination entails, including which vaccines are accepted, poses an additional barrier for travelers – especially those from the global south.

Finally, identifying and tracking the policies themselves is challenging. Countries with COVID- 19 task forces reliably issued updates on dedicated web portals. Others provided more fragmented information based upon the relevant body within their own governments: for instance, entry testing requirements might be available on a nation’s Ministry of Health website, while related flight restrictions would only be found via the Civil Aviation Authority. Countries with more outdated official websites relied heavily on social media to disseminate information, but official documents were easy to miss amid other frequent updates on these channels. For countries with national air carriers, airline websites and social media pages often provide more reliable, comprehensive, and up-to-date access to policy information than government websites. Policies were also rarely available in more than one language, which may pose challenges for interpretation by both travelers and residents who do not speak a country’s official language.

## Conclusion

By the end of 2021, following the observation period, more countries once again followed the response of the United Kingdom by easing targeted restrictions on Southern African nations and continuing a shift to broader enhanced screening. The United States ultimately rescinded its entry ban for travelers from Southern Africa on 31 December, maintaining its 24-hour pre-travel testing requirement even as cases of the variant in the United States soared.[14]

Despite unprecedented numbers of cases, most countries continued to lean on screening measures and tiered “red list” systems. The global shift to a more risk-based approach was in stark contrast to the initial dominance of measures prohibiting travel during the immediate response phase.

Overall, the general confusion and disarray surrounding the implementation of these initial travel restrictions was a disservice to both the travelers impacted and global public health control measures. The challenge of finding up-to-date information amid rapidly changing conditions placed an outside burden on individuals, while the dominant focus policies placed on stopping or delaying importation of a variant that was already globally distributed may have been more effectively directed to domestic preparation efforts. Future mitigation efforts for new emerging variants or other viruses should aim to improve the alignment between response approaches and knowledge about the epidemiology of transmission. As we build the evidence base for what measures are most effective at minimizing the spread of infectious disease across international borders, we hope to see a more risk-based approach to political travel decisions.

## Data Availability

All data produced are available online at covidamp.org

https://covidamp.org/

## Acknowledgements

The authors thank Michael Van Maele for rapidly updating the COVID-AMP website interface to track Omicron-relevant policies and Dr. Alexandra Phelan for informing the legal components of policy data collection.

## Contributorship

RK and EG conceived the analysis and led the research team and effort. JS performed analysis, managed data collection, and drafted manuscript. JS and TW generated figures. JS, MM, SRM, and EW performed data collection. AC, JK, and TW developed data ontology and architecture. All authors reviewed and approved the final draft of the manuscript.

## Declaration of Interests

The authors have no conflict of interest in this work.

## Notes

### Competing Interest Statement

The authors have declared no competing interest.

### Funding Statement

This study received funding from The Rockefeller Foundation, Google Health, the Nuclear Threat Initiative, and Open Philanthropy.

